# Effects of the COVID-19 pandemic on hospital admissions and inpatient mortality in Kenya

**DOI:** 10.1101/2022.10.25.22281489

**Authors:** M Ogero, L Isaaka, L Mumelo, D Kimego, T Njoroge, G Mbevi, C Wanyama, R Lucinde, H Gathuri, M Otiende, C Nzioki, A Wachira, F Mumbi, G Oeri, N Mwangi, R Gitari, D Mugambi, S Namu, A Ithondeka, H Kariuki, Z Kiama, L Mwende, E Jowi, B Muthui, A Kaara, E Sitienei, L Thuranira, I Oginga, J Njagi, E Kamau, E Namulala, G Oketch, O Wandera, S Adhiambo, A Adem, M Ochieng, A Otedo, K Otiende, A Odondi, F Makokha, D Lubanga, J Nyikui, W Masoso, M Manyonge, R Inginia, E Manuthu, D Wafula, C Agutu, R Malangachi, S Biko, Simiyu, J Obare, D Kimutai, B Gituma, J Kyalo, M Timbwa, J Otieno, M Liru, C Nyabinda, S Otieno, R Aman, M Mwangangi, P Amoth, I Were, C Mwangi, K Kasera, W Ng’ang’a, A Tsegaye, C Sherry, B Singa, K Tickell, J Walson, J Berkley, F Were, N Mturi, M Hamaluba, B Tsofa, J Mwangangi, P Bejon, E Barasa, M English, A Nyaguara, EW Kagucia, JAG Scott, S Akech, AO Etyang, A Agweyu

**Affiliations:** KEMRI-Wellcome Trust Research Programme, Nairobi, Kenya; Kenya Paediatric Research Consortium, Nairobi, Kenya; KEMRI-Wellcome Trust Research Programme, Kilifi, Kenya; Department of Health, Machakos County, Kenya; Department of Health, Embu County, Kenya; Department of Health, Nakuru County, Kenya; Department of Health, Nairobi County, Kenya; Department of Health, Kiambu County, Kenya; Department of Health, Busia County, Kenya; Department of Health, Kisumu County, Kenya; Department of Health, Bungoma County, Kenya; Department of Health, Trans Nzoia County, Kenya; Department of Health, Kakamega County, Kenya; Center for Clinical Research, Kenya Medical Research Institute, Nairobi, Kenya; Department of Health, Homabay County, Kenya; Department of Health, Migori County, Kenya; Ministry of Health, Government of Kenya, Nairobi, Kenya; Presidential Policy & Strategy Unit, The Presidency, Government of Kenya, Nairobi, Kenya; Department of Epidemiology, University of Washington, Seattle, Washington, USA; Department of Global Health, University of Washington, Seattle, Washington, USA

## Abstract

**Background:** The impact of COVID-19 in Africa remains poorly defined. We sought to describe trends in hospitalisation due to all medical causes, pneumonia-specific admissions, and inpatient mortality in Kenya before and during the first five waves of the COVID-19 pandemic in Kenya.

**Methods:** We conducted a hospital-based, multi-site, longitudinal observational study of patients admitted to 13 public referral facilities in Kenya from January 2018 to December 2021. The pre-COVID population included patients admitted before 1 March 2020. We fitted time series models to compare observed and predicted trends for each outcome. To estimate the impact of the COVID-19 pandemic, we calculated incidence rate ratios (IRR) and corresponding 95% confidence intervals (CI) from negative binomial mixed-effects models.

**Results:** Out of 302,703 patients hospitalised across the 13 surveillance sites (range 11547 to 57011), 117642 (39%) were admitted to adult wards. Compared with the pre-COVID period, hospitalisations declined markedly among adult (IRR 0.68, 95% CI 0.63 to 0.73) and paediatric (IRR 0.67, 95% CI 0.62 to 0.73) patients. Adjusted in-hospital mortality also declined among both adult (IRR 0.83, 95% CI 0.77 to 0.89) and paediatric (IRR 0.85, 95% CI 0.77 to 0.94) admissions. Pneumonia-specific admissions among adults increased during the pandemic (IRR 1.75, 95% CI 1.18 to 2.59). Paediatric pneumonia cases were lower than pre-pandemic levels in the first year of the pandemic and elevated in late 2021 (IRR 0.78, 95% CI 0.51 to 1.20).

**Conclusions:** Contrary to initial predictions, the COVID-19 pandemic was associated with lower hospitalisation rates and in-hospital mortality, despite increased pneumonia admissions among adults. These trends were sustained after the withdrawal of containment measures that disrupted essential health services, suggesting a role for additional factors that warrant further investigation.

## Introduction

More than two years into the COVID-19 pandemic, questions regarding the toll of COVID-19 in Africa remain unanswered. By the end of 2021, Africa had reported only 7,006 cases per million compared to 36,780 per million people globally[1]. Seroprevalence studies across Africa highlight a wide discrepancy between the high SARS-CoV-2 spread and the burden of observed cases, morbidity and mortality [2-4]. Limited testing capacity may partly explain the under-ascertainment of cases [5]. A more pressing priority is the need for reliable estimates of severe cases and deaths. National routine health information and vital registration systems can offer insights into morbidity and mortality. However, limitations in the completeness of reporting and data quality are often poor [6-8]. The World Health Organisation recommends that countries adapt and strengthen existing hospital-based sentinel surveillance to track the geographical and temporal spread and to assess the severity of the COVID-19 pandemic [9]. Data collected from sentinel surveillance sites can be used to monitor trends and infer the extent of geographical spread. The proportion of confirmed COVID-19 cases among patients hospitalised with acute respiratory infections can also be used as a proxy for severity to estimate the burden of COVID-19 in the population.

Such data may provide an early warning signal of the likelihood of the pandemic to overwhelm the capacity of the healthcare system and guide policy decisions to balance between control measures aimed at curbing the spread of the virus and the collateral societal and economic costs [10]. Kenya reported its first confirmed case of COVID-19 on 13 March 2020. In the subsequent months, the Government announced a series of control measures aimed at curbing the spread of the virus, including restrictions on movement and public gatherings. Since then, the country has experienced five distinct waves: the first two waves peaked in August and November 2020 and were fuelled by the wild-type virus. The Beta and Alpha variants prevailed during the third wave, peaking in April 2021. The fourth wave peaked in August 2021 and was driven by the Delta variant, and the fifth, the Omicron BA1 variant peaking in December 2021 [11]. In this study, we analysed data from a large hospital-based clinical surveillance platform to describe the impact of COVID-19 on hospital admissions and inpatient mortality.

## Methods

### Study design and setting

We undertook a multi-hospital longitudinal study in 13 public hospitals in Kenya. The hospitals are distributed in the densely populated central (highland), western (malaria-endemic) and coastal regions (Figure 1). Among the surveillance sites, 12 are part of a newborn and paediatric Clinical Information Network (CIN) established in 2013 as a partnership between researchers, the Ministry of Health and paediatricians to promote the generation and use of routine data to improve care, understand patient outcomes, and to evaluate interventions [12]. In April 2020, the network activities expanded to include adult medical inpatient surveillance upon request from the Ministry of Health to contribute to the broader national COVID-19 response. At each CIN hospital, one or two full-time clinical officers take responsibility for the coordination of surveillance activities, including providing active feedback to clinical teams on the completion of surveillance forms. One additional surveillance site, Kilifi County Hospital (KCH), is located on the Kenyan Coast and hosts the KEMRI-Wellcome Trust Research Programme. Paediatric and adult inpatient surveillance was established in 1989 and 2007, respectively. Hospital-based surveillance occurs alongside clinical and laboratory research linked to population-based surveillance in the surrounding community [13]. The paediatric department at KCH includes a general ward and a 15-bed high-dependency unit staffed by research clinicians, nurses and data clerks. At the beginning of the pandemic, seven of the 13 surveillance sites were designated COVID-19 treatment centres by the Ministry of Health [14]. The study hospitals are typical of “district-level” health facilities in the region, offering primary care and referral services. High patient numbers and inadequate staffing levels are common, and facilities for advanced critical care are limited.

**Figure 1:**
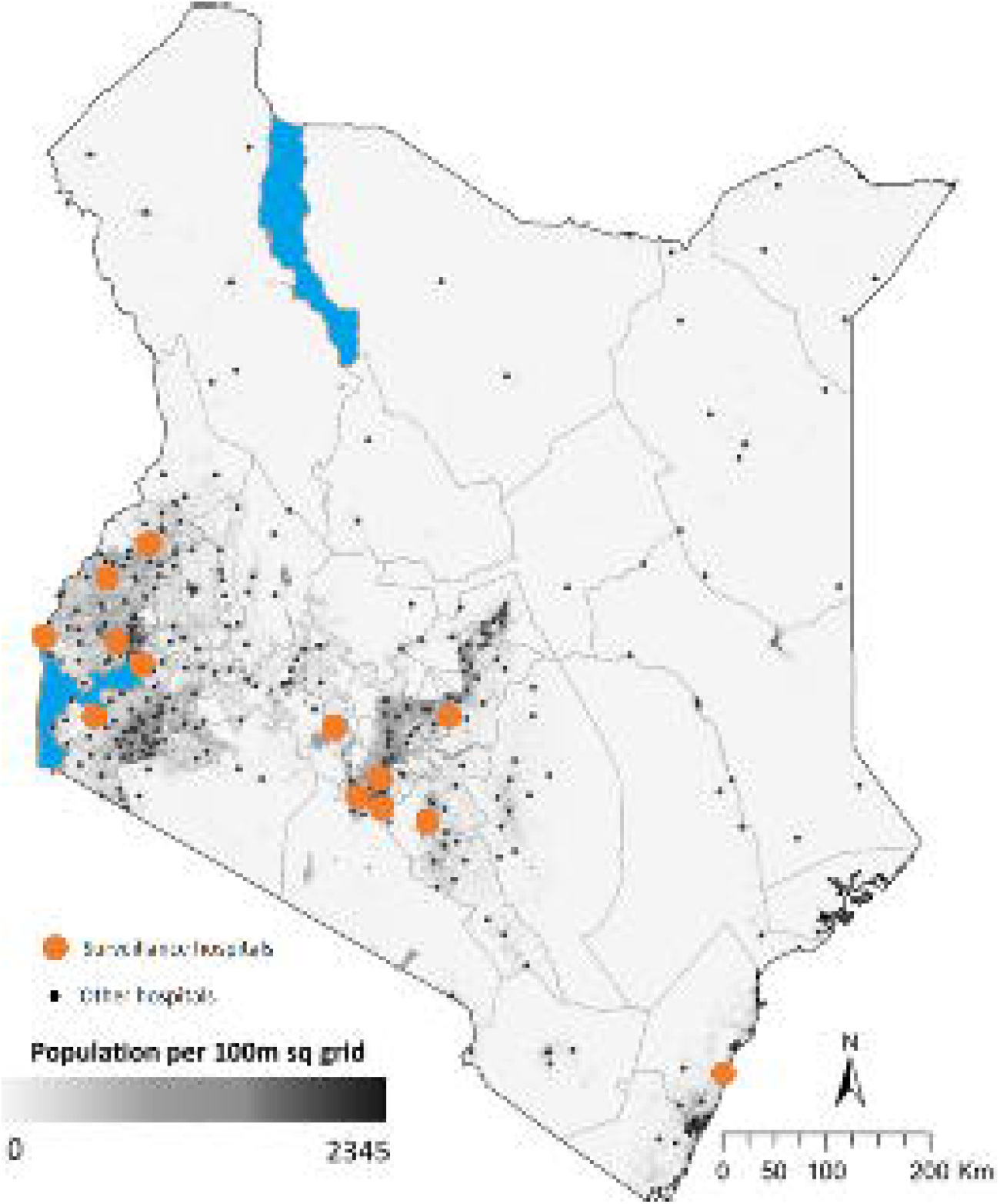
Map of Kenya showing geographical distribution of surveillance hospitals

### Study participants and recruitment

We included all patients discharged from the paediatric and adult medical wards from January 2018 to December 2021. Patients with surgical diagnoses and those admitted to newborn or maternity units were excluded. We used the discharge date to define the inpatient episode rather than the admission date to ensure consistency in populations when reporting outcomes (discharged alive or in-hospital death). The primary aim of the study was to examine the effect of COVID-19 on hospitalisation and inpatient deaths. Admission decisions are made by government-employed clinical officers (non-physician clinicians), who also prescribe initial patient care. Inpatient management is typically delivered by a team of medical officers and medical officer interns, and nurses under the supervision of one or more specialist physicians. Scheduled medical ward rounds are conducted daily during admission with concurrent documentation of observations and updating clinical plans and diagnoses in the inpatient medical records until discharge. In December 2020 and January 2021, a nationwide strike led to the suspension of inpatient services in public health facilities, including all 13 surveillance hospitals.

### Data collection

The methods of data collection and analysis in CIN and KCH are reported elsewhere [15-17]. In brief, the hospitals implement structured data collection tools (admission and discharge form). The tools capture data on patient clinical characteristics, basic laboratory tests conducted at admission diagnoses assigned, treatments prescribed, and clinical outcomes. The study did not provide direct resources to support RT-PCR testing for COVID-19 but instead relied on the available capacity at the hospitals and the counties. A diagnosis of suspected COVID-19 was assigned based on the national case definition: severe acute respiratory illness (fever or cough or shortness of breath; AND requiring hospitalisation) AND in the absence of an alternative diagnosis that fully explains the clinical presentation[18]. Confirmed COVID-19 was defined by a positive real-time reverse transcription-polymerase chain reaction (RT-PCR) for severe acute respiratory syndrome coronavirus 2 (SARS-CoV-2) in a respiratory sample at any time during or before admission (Supplementary Table 1). Data were abstracted on-site from inpatient paper records into an electronic tool following admission (KCH and adult CIN) and at discharge by trained staff using standard operating procedures. Data for adults admitted to CIN hospitals before April 2020 were extracted retrospectively from archived medical records. Daily bed return entries were used to cross-validate the total number of monthly discharges and deaths. Where a discrepancy was found between the sources, the higher of the two was captured. The electronic data capture tool has built-in soft and hard error validation. De-identified data from all hospitals are synchronised daily to KEMRI central servers. Centrally, additional quality checks are performed, allowing the supervisory team to provide feedback to resolve discrepancies identified with individual sites or collectively through weekly data review meetings.

### Statistical methods

We included all eligible admissions from the population of hospitalised patients at the surveillance sites. Therefore, no formal sample size estimation was conducted.

Data were assembled across 13 hospitals for patients admitted to paediatric and adult wards from 1 January 2018 to 31 December 2021. We excluded patients admitted during the healthcare workers’ strike in December 2020 and January 2021. Our primary exposure was the period of admission grouped as pre-covid for the patients admitted before 1 March 2020 and during the COVID-19 pandemic for the patients admitted at any time from 1 March 2020 to 31 December 2021.

### Statistical analyses

We examined the impact of the COVID-19 pandemic on the following outcomes 1) adult admissions, 2) paediatric admissions, 3) adult deaths, 4) paediatric deaths, 5) adult pneumonia admissions, and 6) paediatric pneumonia admissions. For each outcome, we analysed trends over the study period and estimated the effect of the pandemic.

To study trends, data from patients admitted from 1 January 2018 to 31 December 2019 were used to train a time series model with adjustments for seasonality and hospital identity. The resultant model was then used to forecast monthly counts and compute corresponding 95% prediction intervals for those admitted from 1 January 2020 to 31 December 2021. We compared the observed and predicted numbers visually via line charts. The *forecast* R package was used for this analysis.

We estimated the impact of the COVID-19 pandemic using negative binomial mixed-effects models (with hospital identity as a random effect) to account for over-dispersion of the monthly counts of each outcome. We specified the correlation structure in the model using autoregressive (AR) and moving average (MA) parameters, which were determined by examining the autocorrelation factor (ACF) and partial autocorrelation factor (PACF) plots. Models were fitted using the *glmmPQL* function available in MASS (Modern Applied Statistics with S) package [19]. We reported the results as adjusted incidence rate ratios and corresponding 95% confidence intervals and p-values. All analyses were performed using R statistical programming language version 3.6.3 [20].

Kenya Medical Research Institute (KEMRI) Scientific and Ethics Review Unit approved the study.

## Results

Between January 2018 and December 2021, 302,703 patients were admitted for care across the 13 hospitals (range 11547 to 57011). Patients included 148,318 (49.2%) aged under five years, and 117,642 (39%) admitted to the adult wards. Overall, 21,227 (13.8%) died while receiving inpatient care, ranging across hospitals from 906/9467 (9.6%) to 1546/7453 (20.7%) (Table 1).

**Table 1:** Hospital and patient population characteristics

| Characteristics | H1 | H2 | H3 | H4 | H5 | H6 | H7 | H8 | H9 | H10 | H11 | H12 | H13 | All sites |
| --- | --- | --- | --- | --- | --- | --- | --- | --- | --- | --- | --- | --- | --- | --- |
| Provides ICU care* | Yes | No | Yes | No | Yes | Yes | No | No | Yes | No | No | No | No |  |
| Designated COVID-19 treatment facility | No | No | Yes | No | Yes | Yes | Yes | No | Yes | No | Yes | Yes | No |  |
| Total inpatients | 19227 | 27785 | 21213 | 11547 | 22657 | 28628 | 57011 | 17238 | 26012 | 15711 | 23173 | 15036 | 17462 | 302703 |
| Paediatric wards < 5 years (%) | 6370<br>(33.6%) | 12095<br>(43.6%) | 12123<br>(57.3%) | 2390<br>(20.7%) | 11783<br>(52.4%) | 18716<br>(65.9%) | 22988<br>(40.4%) | 7938<br>(46.3%) | 11606<br>(44.7%) | 9303<br>(59.3%) | 19374<br>(84.4%) | 9990<br>(66.8%) | 3642<br>(20.9%) | 148318<br>(49.2%) |
| Paediatric wards >5 years (%) | 2192<br>(11.6%) | 5836<br>(21%) | 1346<br>(6.4%) | 769<br>(6.7%) | 4998<br>(22.2%) | 1767<br>(6.2%) | 6246<br>(11%) | 2876<br>(16.8%) | 3798<br>(14.6%) | 1183<br>(7.5%) | 2336<br>(10.2%) | 1446<br>(9.7%) | 640<br>(3.7%) | 35433<br>(11.8%) |
| Adult wards (%) | 10381<br>(54.8%) | 9820<br>(35.4%) | 7697<br>(36.4%) | 8384<br>(72.6%) | 5724<br>(25.4%) | 7925<br>(27.9%) | 27708<br>(48.7%) | 6341<br>(37%) | 10553<br>(40.7%) | 5202<br>(33.2%) | 1252<br>(5.5%) | 3509<br>(23.5%) | 13146<br>(75.4%) | 117642<br>(39%) |
| Crude mortality (%) | 2365<br>(12.3%) | 2843<br>(10.2%) | 3222<br>(15.2%) | 2037<br>(17.6%) | 2786<br>(12.3%) | 3689<br>(12.9%) | 6474<br>(11.4%) | 1604<br>(9.3%) | 3282<br>(12.6%) | 2321<br>(14.8%) | 2007<br>(8.7%) | 2334<br>(15.5%) | 1873<br>(10.7%) | 36837<br>(12.2%) |

\*In March 2021

### Trends in all-cause hospitalisation

Among adults, the variation in hospitalisation showed seasonal variation over the pre-pandemic period. In March 2020, the number of patients hospitalised across the hospitals fell sharply and remained lower than predicted levels until March 2021, when there was a transient rebound to baseline levels. Thereafter, numbers fell below the lower predicted limit until September 2021. From September 2021 to December 2021, adult hospitalisation numbers remained within the predicted bounds. (Figure 2). Overall, we observed a 32% decline in adult hospitalisation numbers during the pandemic period up to December 2021 compared to the pre-pandemic period (IRR 0.68, 95% CI 0.63 to 0.73; P<0.001) (Table 2).

**Table 2:**
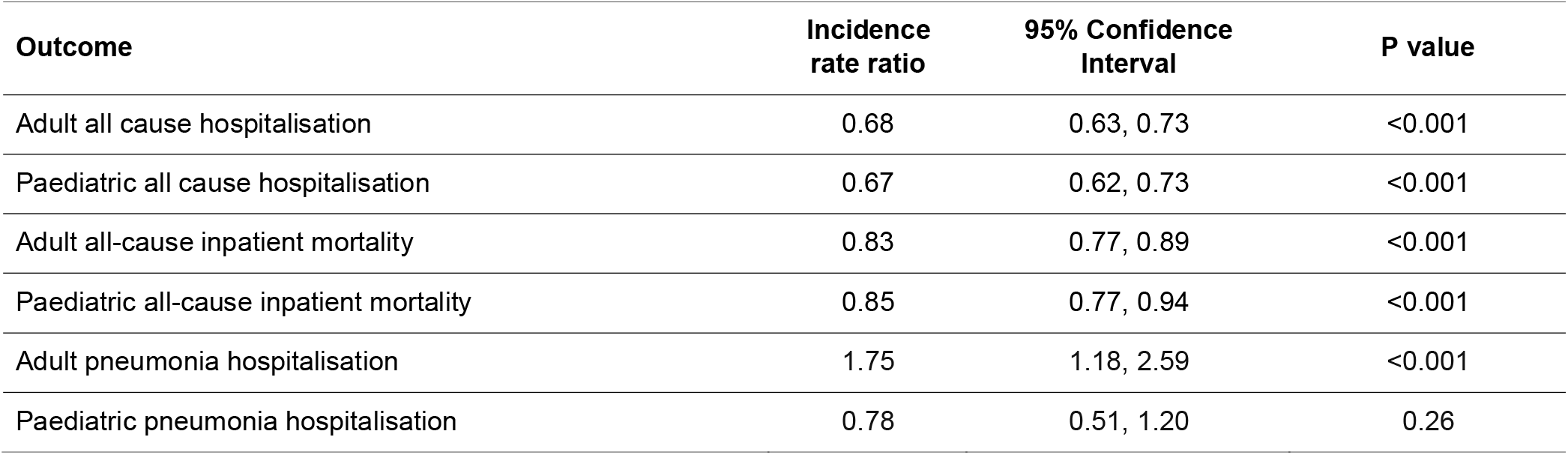
Adjusted incidence rate ratios for associations between study outcomes and the COVID-19 pandemic

**Figure 2:**
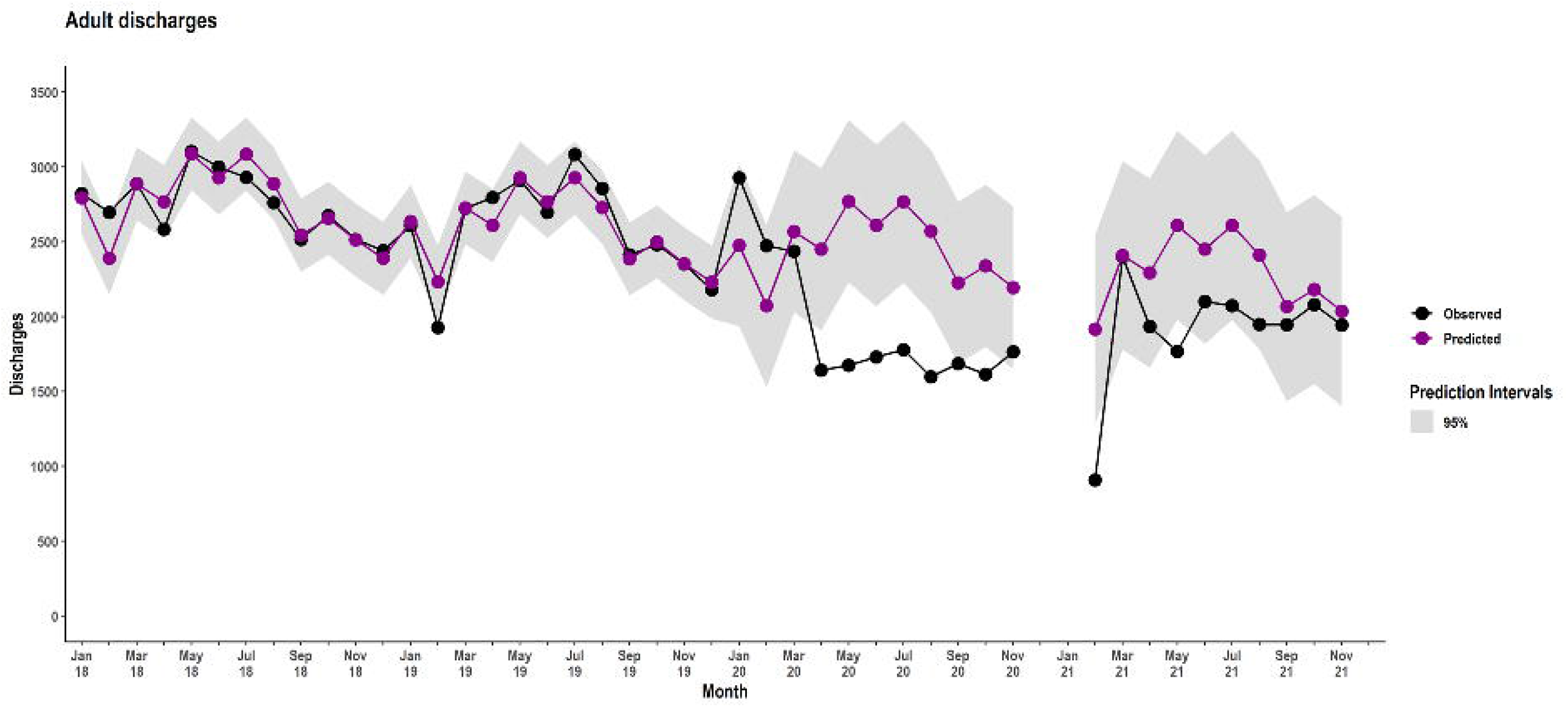
Trends in adult all-cause hospitalisation

Paediatric admissions exceeded the upper bound of the predicted limit marginally in February 2020, returning to predicted levels in subsequent months. Similar to the trend among adults, a decline was observed from April 2020 that was sustained until March 2021. There was a second decrease between April and August 2021 (Figure 3). Paediatric hospitalisation numbers reduced by 33% during the pandemic (IRR 0.67, 95% CI 0.62 to 0.73; P<0.001) (Table 2).

**Figure 3:**
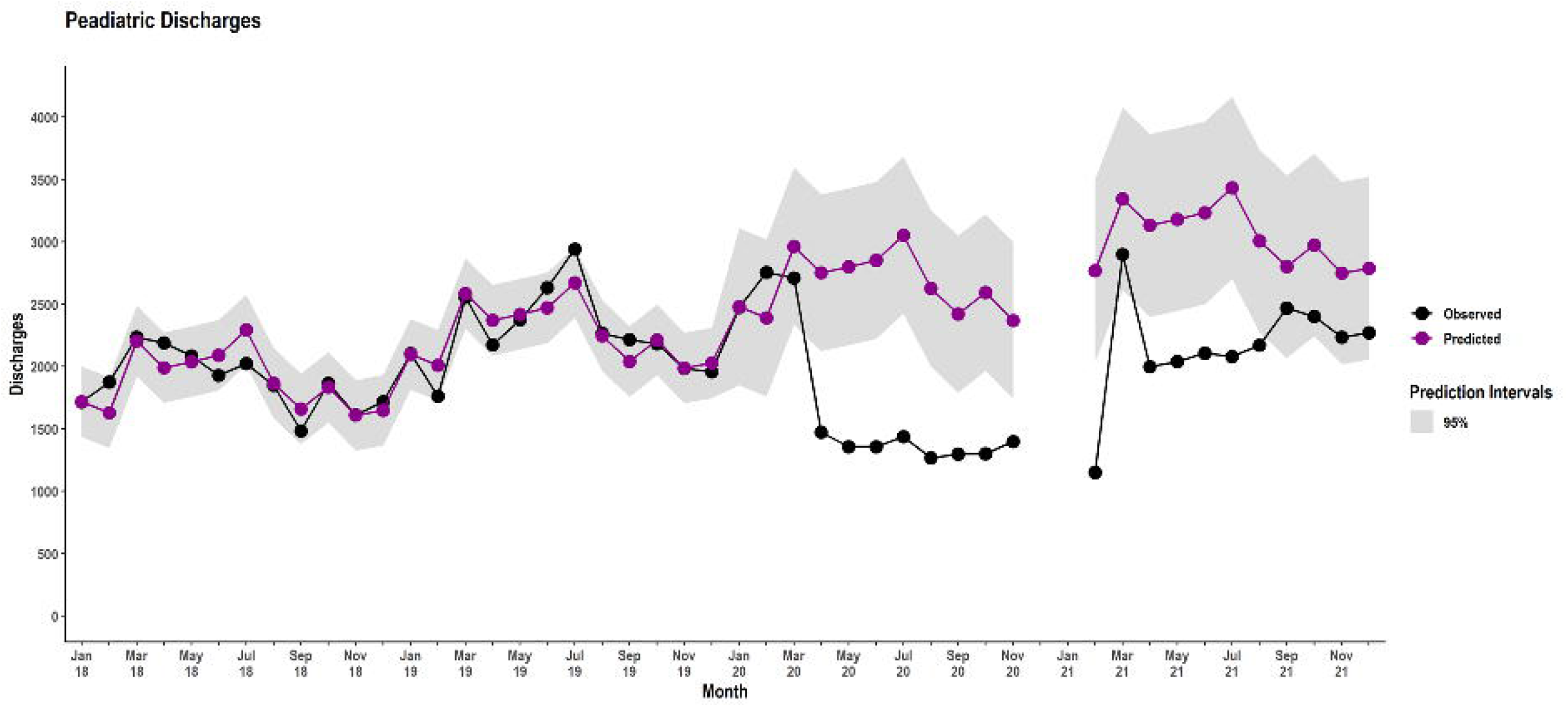
Trends in paediatric all-cause hospitalisation

### Trends in inpatient mortality

Adult inpatient mortality reduced in April 2020 but remained within predicted limits until March 2021, after which the observed deaths rebounded but did not exceed the upper predicted limit (Figure 4). Adult all-cause inpatient mortality reduced by 17% during the pandemic (IRR 0.83, 95% CI 0.77 to 0.89; P<0.001) (Table 2).

**Figure 4:**
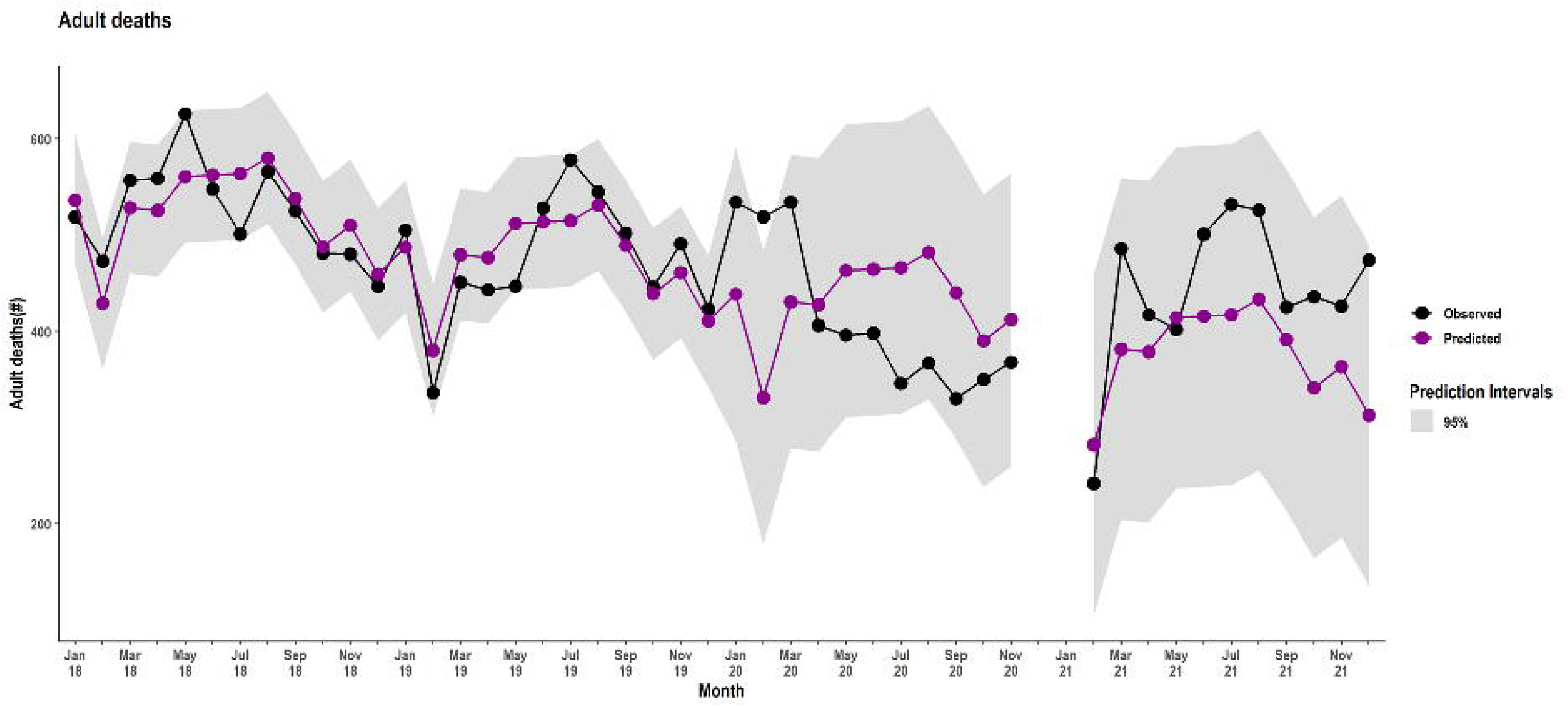
Trends in adult all-cause inpatient mortality

Paediatric inpatient mortality declined in April 2020, falling below the predicted lower limit in June 2020. Mortality remained low, transiently rising in March 2021, dropping below baseline limits again in subsequent months, and rising to predicted levels from September to December 2021 (Figure 5). Inpatient paediatric mortality was significantly lower during the pandemic compared to the study interval before the pandemic (IRR 0.85, 95% CI 0.77 to 0.94; P<0.001) (Table 2).

**Figure 5:**
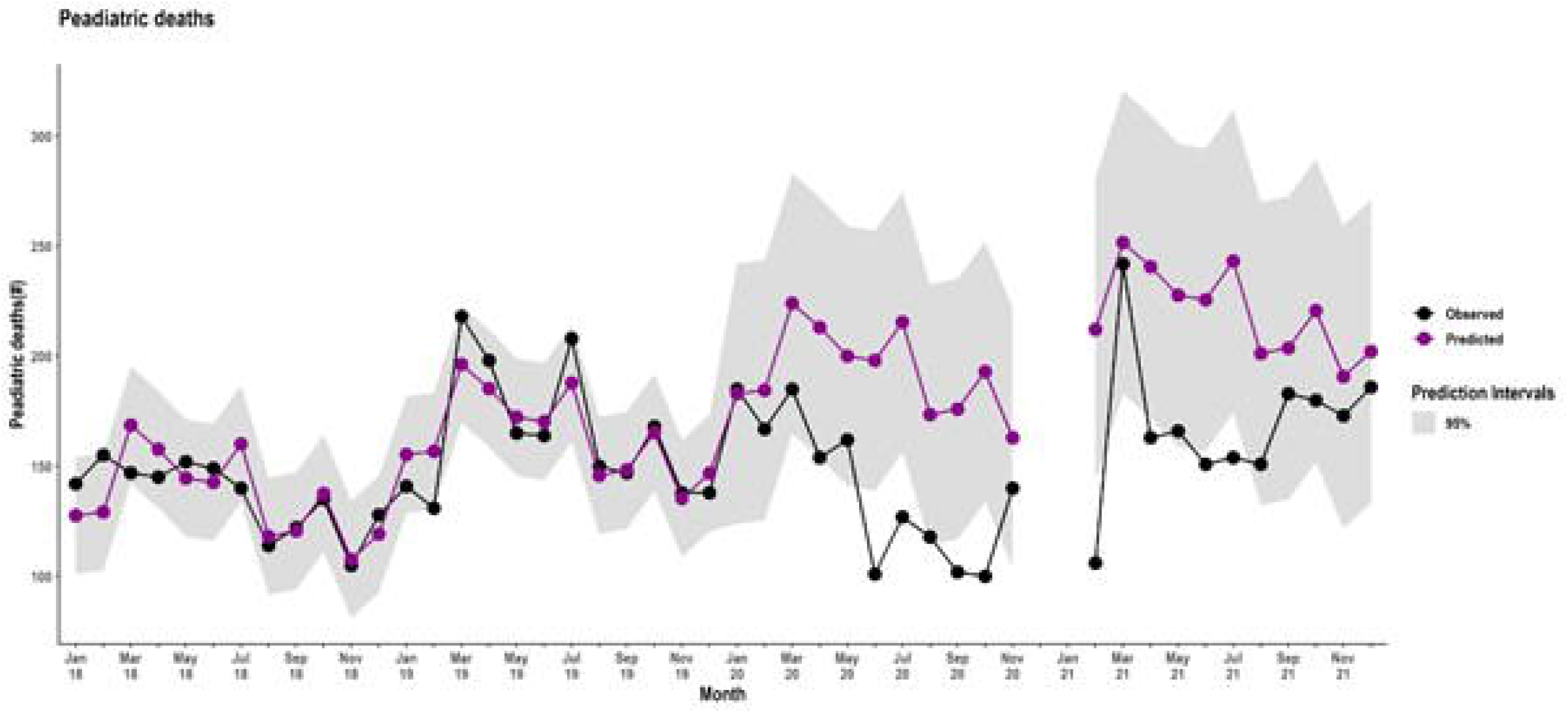
Trends in paediatric all-cause inpatient mortality

### Trends in pneumonia hospitalisation

Adult pneumonia increased in February 2020 and remained above the predicted levels throughout the pandemic, peaking in March and April 2021 (Figure 6). Adult pneumonia admissions increased by 75% during the pandemic (IRR 1.75, 95% CI 1.18 to 2.59; P<0.001) (Table 2).

**Figure 6:**
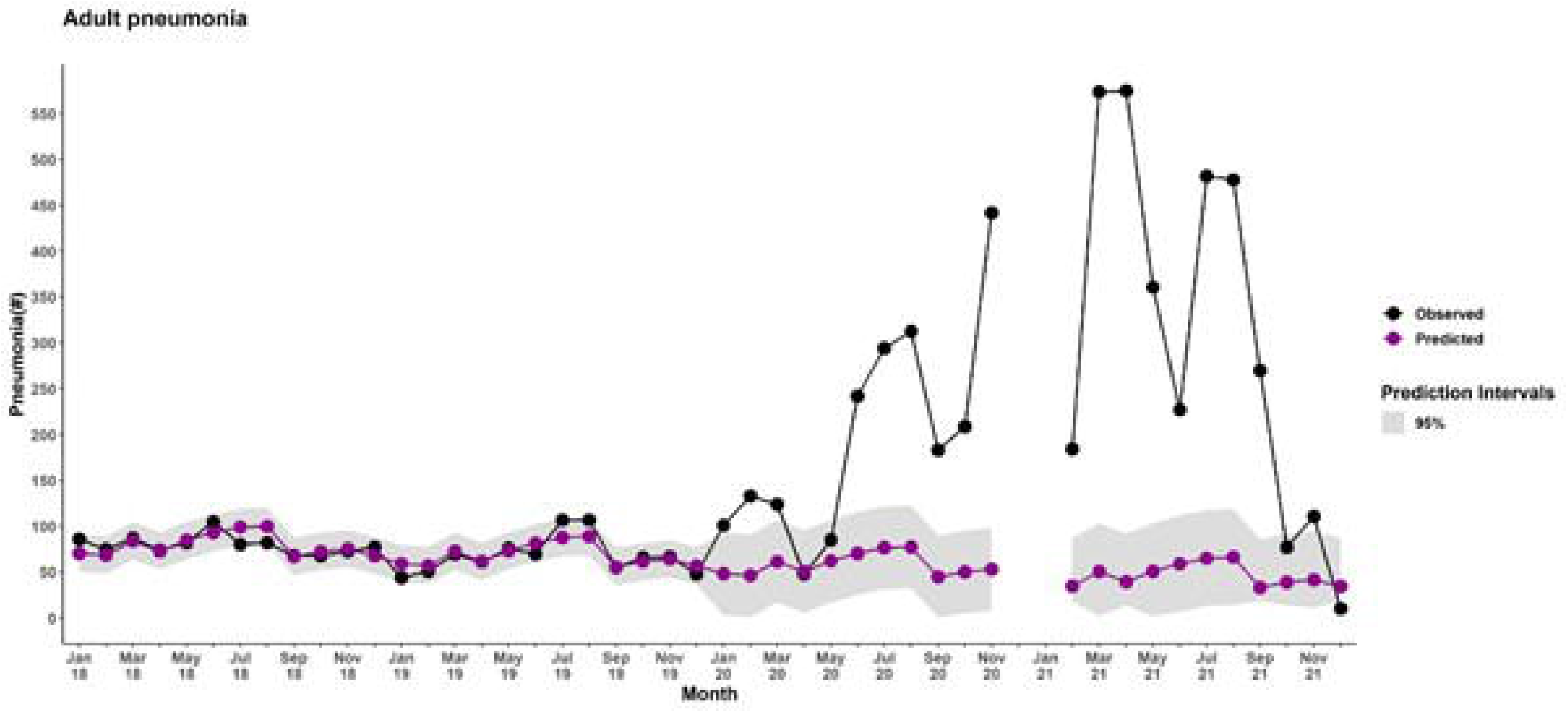
Trends in adult pneumonia hospitalisation

There was a reduction in paediatric pneumonia admissions from the beginning of the pandemic in April 2020 until the end of the 2020. The admission numbers rose in early 2021, exceeding the upper predicted limits in March and again from August to December 2021 (Figure 7). The net effect of the pandemic on paediatric pneumonia admissions was not statistically significant (IRR 0.78, 95% CI 0.51 to 1.20; P=0.26) (Table 2).

**Figure 7:**
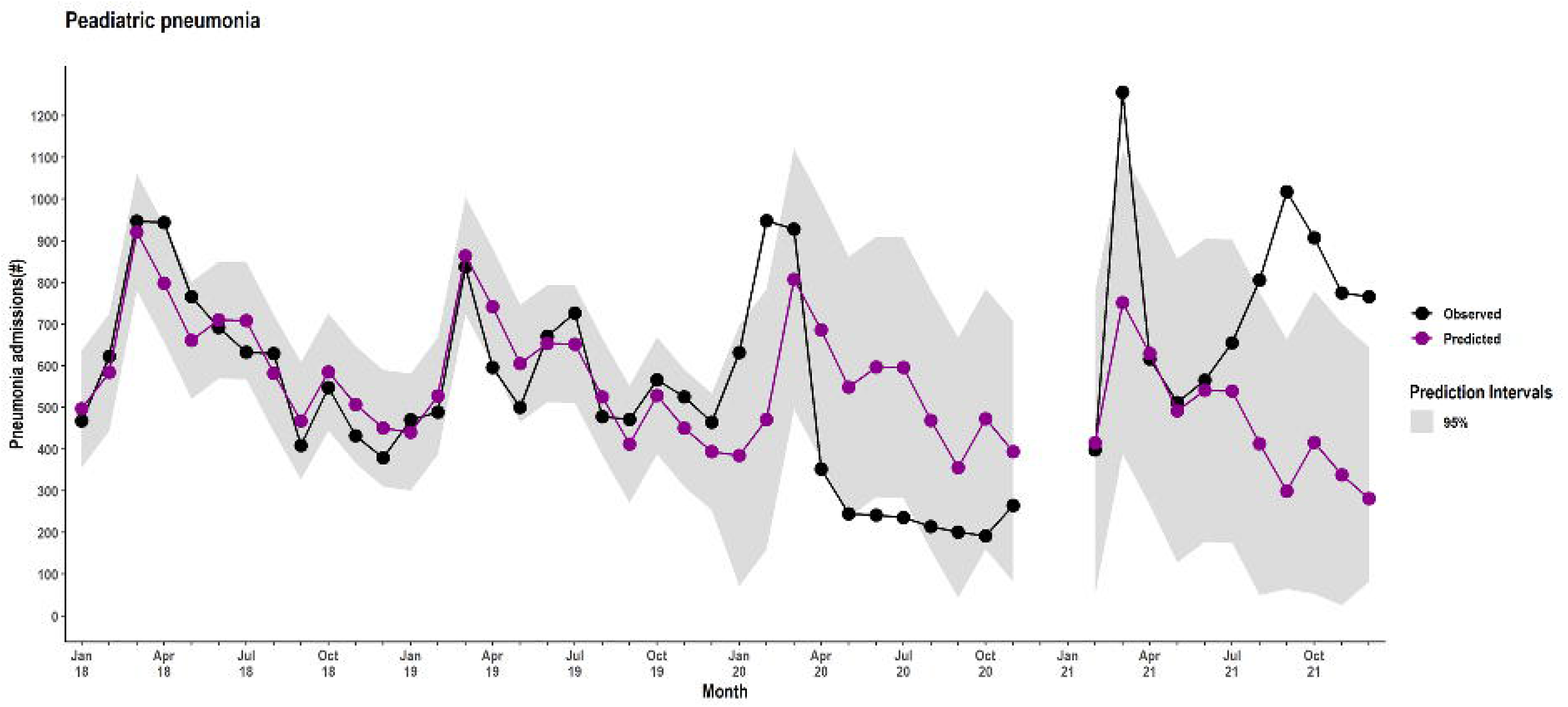
Trends in paediatric pneumonia hospitalisation

## Discussion

In this four-year longitudinal analysis, we observed substantial reductions in all-cause inpatient mortality and hospitalisation among adults and children during the pandemic. These results contradict initial predictions of increased strain on hospital systems [21, 22]. We postulate several possibilities underlying these trends.

Although healthcare providers and patients requiring emergency health services were exempt from the lockdowns, various studies have demonstrated reduced utilisation of outpatient health services during the pandemic [23-26]. It is plausible that the movement restrictions limited access to transport, and the perceived increased risk of contracting COVID-19 in hospital settings led to reduced or delayed care seeking [27, 28]. However, published studies have reported a return to normal levels of service utilisation by early 2021 [23-25]. Our analysis showed a brief rebound in admission and inpatient mortality around March 2021 and a subsequent decline in paediatric and adult admissions and paediatric deaths. Interestingly, adult mortality peaked in July and August 2021, coinciding with the peak of the Delta wave. This finding is consistent with results from a population-based study in coastal Kenya which found excess mortality among individuals aged 45-64 years and ≥65 years only during July 2021 [29].

Like many countries in the region, Kenya has faced challenges with testing for SARS-CoV-2 [5]. Therefore, an alternative explanation for the unexpectedly low hospitalisation and mortality observed may be low overall case numbers in the Kenyan population. However, seroprevalence studies suggest progressive dissemination of the pandemic in the Kenyan population beginning in early 2020 [30], approaching 50% among blood donors by March 2021 [31], rising to 85% among unvaccinated individuals in the general population in early 2022 [32]. The young population structure in Africa, with a predominance of asymptomatic and mild presentations of infection, may offset the high burden of other risk factors for poor outcomes in the region [33, 34] and partly explain the lower occurrence of severe and fatal infections in sub-Saharan Africa [35-37].

Limited data from population-based surveillance has ignited speculation about a hidden burden of severe COVID-19 cases and deaths in the community. A cross-sectional descriptive postmortem study conducted in a tertiary hospital in Zambia between June and September 2020 confirmed SARS-CoV-2 infection in 70/364 (19.2%) participants, 51 (73%) of whom died in the community. However, without contemporaneous data from previous years to demonstrate the additional contribution of COVID-19 to mortality, the diagnosis may have been an incidental finding at postmortem in an urban setting at the peak of the pandemic [38]. In our study, the similarities in trends between adults and children (among whom the burden of severe and fatal COVID-19 is much less common) may provide insights that distinguish between morbidity and mortality due to COVID-19 and the indirect effects of the pandemic on health service utilisation. There is also emerging reliable evidence from Health and Demographic Surveillance sites suggesting that mortality in Africa may be considerably lower than initially projected [29, 39].

There was a sharp increase in adult admissions due to pneumonia without corresponding changes in inpatient mortality. We speculate that this trend may have been the result of a reduced threshold for admission of adult patients with acute respiratory illness during the pandemic out of caution or to allow for confirmatory diagnosis of COVID-19. Notably, the lowest number of recorded adult pneumonia admissions coincided with the fifth pandemic wave driven by the milder Omicron variant. Among children, the decline in pneumonia admissions may have been the result of the aggressive implementation of COVID-19 preventive measures, including school closures, improved hand hygiene, social distancing, and wearing masks [40]. This theory is supported by the resurgence in cases in 2021 after schools reopened and the relaxation of other control interventions.

The strengths of this study include the multi-centre longitudinal design with robust quality assurance for the administrative data analysed. To our knowledge, this is the first study of this scale in sub-Saharan Africa describing trends in all-cause hospital outcomes COVID-19 for both adults and children.

The study had several limitations. First, unmeasured variables are likely to have contributed to the findings. Therefore, the results should be interpreted as associations rather than causal. Although the surveillance sites are located in densely populated regions of the country, they are not a representative sample of hospitals in Kenya. However, data captured in the national health information system that includes all registered health facilities in the country did not reveal a noticeable increase in admissions coinciding with the pandemic period (Supplementary Table 2). The number of hospitalised cases of severe acute respiratory infections may indicate the disease burden in the population. However, because of the coincident introduction of pandemic control measures that may have affected care-seeking, this assumption can only be validated with carefully designed community-based surveys on morbidity and mortality.

We relied on the health system capacity for SARS-CoV-2 testing. Almost one-third of adult patients hospitalised during the pandemic fulfilled the case definition for suspected COVID-19, and disease was only confirmed in approximately one-third of those tested (Supplementary Table 1). Finer age stratification would have allowed us to explore age-specific trends in outcomes and improve risk adjustment in our models. Indeed, the excess mortality analysis from a population-based surveillance platform in coastal Kenya revealed significant excess mortality only among residents aged 65 years and above [29]. Unfortunately, these data were unavailable due to variable documentation in the pre-pandemic adult dataset. The absence of a control population and population denominators further precludes attribution.

Kenya received its first COVID-19 vaccines on 3 March 2021. At present, the vaccination coverage, is progressively increasing with broader availability. However, by 31 December 2021, the national COVID-19 vaccination programme had reached only 15% of the adult population with wide geographic disparities. Therefore, vaccination status is unlikely to have contributed materially to the findings. We are also unable to predict the effects of future waves of the pandemic that may be driven by SARS-CoV-2 variants with immune escape properties. This observation has important implications for vulnerable populations that remain susceptible to infection in the face of potential future waves of highly transmissible variants. It signals the urgency to refocus the current vaccination strategy towards the rural-dwelling elderly, a strategy that is backed by evidence from cost-effectiveness analyses [41].

## Conclusions

We harnessed a surveillance platform embedded within a Learning Health System to demonstrate lower hospitalisation rates and in-hospital mortality, despite increased pneumonia admissions among adults. These trends were sustained after the withdrawal of containment measures that resulted in the disruption of essential health services, suggesting a role for additional factors that warrant further investigation. Pneumonia admissions among adults remained elevated, while paediatric pneumonia cases declined in the first year of the pandemic and increased subsequently after the relaxation of government-mandated control measures.

## Data Availability

All data produced in the present study are available upon reasonable request to the authors

## Declarations

### Ethics approval and consent to participate

This Scientific and Ethical Review Committee of the Kenya Medical Research Institute (approval number: KEMRI/SERU/CGMR-C/203/4085), the Oxford Tropical Research Ethics Committee (44-20), and the London School of Hygiene and Tropical Medicine Research Ethics Committee (26950) gave ethical approval for this work. Additionally, the study was approved by the Ministry of Health, and participating counties, with the Medical Superintendents of hospitals giving consent for participation. Individual consent for access to de-identified patient data was not required.

### Data Availability

Data are available upon reasonable request.

For the purpose of Open Access, the author has applied a CC-BY public copyright license to any accepted manuscript version arising from this submission.

## Acknowledgements

We are grateful to the hospital teams, including staff, patients and caregivers, and Ministry of Health colleagues, for supporting the Clinical Information Network and all clinical surveillance activities of the Kilifi HDSS. This article is published with the permission of the Director of the Kenya Medical Research Institute.

