## Supplementary Table 1 for "Effects of the COVID-19 pandemic on hospital admissions and inpatient mortality in Kenya"

**Supplementary Table 1: COVID-19 surveillance among SARI cases in COVID-19 treatment centres from April-Dec 2020**

|  | **H5** | **H6** | **H8** | **H10** | **H12** | **H13** |
| --- | --- | --- | --- | --- | --- | --- |
| **Total** | 3152 | 5396 | 3885 | 1933 | 1298 | 6103 |
| COVID-19 tested – positive (%) | 214 (6.8%) | 399 (7.4%) | 30 (0.8%) | 194 (10%) | 262 (20.2%) | 116 (1.9%) |
| COVID-19 tested – Negative (%) | 178 (5.6%) | 679 (12.6%) | 44 (1.1%) | 410 (21.2%) | 71 (5.5%) | 259 (4.2%) |
| SARI cases | 1041 | 2829 | 896 | 700 | 782 | 1337 |
| COVID-19 suspected - not tested (%) | 650 (62.4%) | 1752 (61.9%) | 823 (91.9%) | 101 (14.4%) | 449 (57.4%) | 963 (72%) |
