## Supplementary Table 2 for "Effects of the COVID-19 pandemic on hospital admissions and inpatient mortality in Kenya"

**Supplementary Table 2: Monthly case fatality ratios for all surveillance hospitals and Kenya Health Information System from January 2018- December 2021**

Supplementary Table 2.1: Surveillance hospital monthly adult medical inpatient case fatality ratios

| **Year** | **Jan** | **Feb** | **Mar** | **Apr** | **May** | **Jun** | **Jul** | **Aug** | **Sep** | **Oct** | **Nov** | **Dec** |
| --- | --- | --- | --- | --- | --- | --- | --- | --- | --- | --- | --- | --- |
| 2,018 | 519/2822 (18.4%) | 473/2697 (17.5%) | 557/2889 (19.3%) | 559/2584 (21.6%) | 626/3104 (20.2%) | 548/3001 (18.3%) | 501/2931 (17.1%) | 566/2762 (20.5%) | 525/2517 (20.9%) | 481/2677 (18%) | 480/2516 (19.1%) | 447/2442 (18.3%) |
| 2,019 | 505/2609 (19.4%) | 336/1928 (17.4%) | 451/2725 (16.6%) | 443/2796 (15.8%) | 447/2913 (15.3%) | 528/2696 (19.6%) | 578/3084 (18.7%) | 545/2857 (19.1%) | 502/2415 (20.8%) | 446/2481 (18%) | 491/2351 (20.9%) | 423/2179 (19.4%) |
| 2,020 | 534/2929 (18.2%) | 519/2474 (21%) | 534/2437 (21.9%) | 406/1642 (24.7%) | 396/1674 (23.7%) | 398/1731 (23%) | 346/1779 (19.4%) | 367/1598 (23%) | 330/1686 (19.6%) | 350/1614 (21.7%) | 368/1764 (20.9%) | 140/990 (14.1%) |
| 2,021 | 102/482(21.2%) | 242/907(26.7%) | 486/2400(20.2%) | 417/1934(21.6%) | 402/1768(22.7%) | 501/2101(23.8%) | 532/2073(25.7%) | 526/1948(27%) | 425/1945(21.9%) | 436/2080(21%) | 426/1944(21.9%) | 474/1829(25.9%) |

Supplementary Table 2.2: Surveillance hospital monthly paediatric medical inpatient case fatality ratios

| **Year** | **Jan** | **Feb** | **Mar** | **Apr** | **May** | **Jun** | **Jul** | **Aug** | **Sep** | **Oct** | **Nov** | **Dec** |
| --- | --- | --- | --- | --- | --- | --- | --- | --- | --- | --- | --- | --- |
| 2,018 | 142/1714 (8.3%) | 155/1876 (8.3%) | 147/2234 (6.6%) | 145/2190 (6.6%) | 152/2085 (7.3%) | 149/1929 (7.7%) | 140/2023 (6.9%) | 114/1847 (6.2%) | 122/1485 (8.2%) | 135/1866 (7.2%) | 105/1613 (6.5%) | 128/1718 (7.5%) |
| 2,019 | 141/2103(6.7%) | 131/1762 (7.4%) | 218/2554 (8.5%) | 198/2173 (9.1%) | 165/2373 (7%) | 164/2634 (6.2%) | 208/2942 (7.1%) | 150/2265 (6.6%) | 147/2216 (6.6%) | 168/2181 (7.7%) | 138/1984 (7%) | 138/1958 (7%) |
| 2,020 | 185/2474 (7.5%) | 167/2755 (6.1%) | 185/2710 (6.8%) | 154/1474 (10.4%) | 162/1356 (11.9%) | 101/1355 (7.5%) | 127/1438 (8.8%) | 118/1266 (9.3%) | 102/1298 (7.9%) | 100/1301 (7.7%) | 140/1398 (10%) | 43/698 (6.2%) |
| 2,021 | 45/350 (12.9%) | 106/1151 (9.2%) | 242/2899 (8.3%) | 163/1999 (8.2%) | 166/2040 (8.1%) | 151/2108 (7.2%) | 154/2080 (7.4%) | 151/2171 (7%) | 183/2468 (7.4%) | 180/2401 (7.5%) | 173/2234 (7.7%) | 186/2270 (8.2%) |

Supplementary Table 2.3: Kenya Health Information System adult medical inpatient case fatality ratios

| **Year** | **Jan** | **Feb** | **Mar** | **Apr** | **May** | **Jun** | **Jul** | **Aug** | **Sep** | **Oct** | **Nov** | **Dec** |
| --- | --- | --- | --- | --- | --- | --- | --- | --- | --- | --- | --- | --- |
| 2,018 | 4292/60293 (7.1%) | 2705/54972 (4.9%) | 3074/60769 (5.1%) | 3123/61285 (5.1%) | 3097/66787 (4.6%) | 3048/64823 (4.7%) | 3553/68955 (5.2%) | 3060/59761 (5.1%) | 2839/60013 (4.7%) | 2777/61786 (4.5%) | 2645/61666 (4.3%) | 2598/56147 (4.6%) |
| 2,019 | 3139/67371 (4.7%) | 2588/59477 (4.4%) | 3267/71208 (4.6%) | 2975/68093 (4.4%) | 3161/80091 (3.9%) | 3484/76649 (4.5%) | 3893/87998 (4.4%) | 3256/77113 (4.2%) | 3117/74253 (4.2%) | 3042/72927 (4.2%) | 3113/67023 (4.6%) | 3119/67383 (4.6%) |
| 2,020 | 3554/82776 (4.3%) | 3389/78180 (4.3%) | 2950/75268 (3.9%) | 2792/51625 (5.4%) | 2787/57475 (4.8%) | 3072/63011 (4.9%) | 3348/68420 (4.9%) | 3328/65715 (5.1%) | 3305/67592 (4.9%) | 3555/76026 (4.7%) | 3512/72705 (4.8%) | 2988/64885 (4.6%) |
| 2,021 | 2834/67585 (4.2%) | 3074/69260 (4.4%) | 4051/80417 (5%) | 3697/72574 (5.1%) | 3762/78355 (4.8%) | 3695/82130 (4.5%) | 3992/74850 (5.3%) | 4303/78569 (5.5%) | 3690/77491 (4.8%) | 4033/74285 (5.4%) | 3228/77489 (4.2%) | 3566/80681 (4.4%) |

Supplementary Table 2.4: Kenya Health Information System paediatric medical inpatient case fatality ratios

| **Year** | **Jan** | **Feb** | **Mar** | **Apr** | **May** | **Jun** | **Jul** | **Aug** | **Sep** | **Oct** | **Nov** | **Dec** |
| --- | --- | --- | --- | --- | --- | --- | --- | --- | --- | --- | --- | --- |
| 2,018 | 738/21535 (3.4%) | 634/23860 (2.7%) | 998/25300 (3.9%) | 664/25429 (2.6%) | 646/25773 (2.5%) | 661/26227 (2.5%) | 754/30752 (2.5%) | 509/21708 (2.3%) | 503/20639 (2.4%) | 545/22560 (2.4%) | 529/20460 (2.6%) | 515/19375 (2.7%) |
| 2,019 | 617/24364 (2.5%) | 666/26107 (2.6%) | 765/28888 (2.6%) | 916/27106 (3.4%) | 757/28702 (2.6%) | 964/30658 (3.1%) | 822/33053 (2.5%) | 842/29821 (2.8%) | 618/23959 (2.6%) | 597/23673 (2.5%) | 631/23554 (2.7%) | 602/22387 (2.7%) |
| 2,020 | 815/27307 (3%) | 729/29660 (2.5%) | 725/27372 (2.6%) | 642/14383 (4.5%) | 545/14636 (3.7%) | 548/15675 (3.5%) | 571/15998 (3.6%) | 574/15576 (3.7%) | 614/16496 (3.7%) | 553/17187 (3.2%) | 610/16085 (3.8%) | 460/15161 (3%) |
| 2,021 | 527/16133 (3.3%) | 618/20994 (2.9%) | 1042/30199 (3.5%) | 614/20941 (2.9%) | 566/21433 (2.6%) | 621/23232 (2.7%) | 710/24084 (2.9%) | 735/27073 (2.7%) | 670/28923 (2.3%) | 1265/26604 (4.8%) | 640/26510 (2.4%) | 756/26957 (2.8%) |
